# Feasibility of Real-Time Cognitive Load Assessment Using Electronic Health Record Data and Wearable Sensors in Emergency Medicine: A Pilot Study

**DOI:** 10.1101/2025.06.02.25328813

**Authors:** David A. Kim, Carl Preiksaitis, Christian Rose

**Affiliations:** Department of Emergency Medicine, Stanford University School of Medicine, Palo Alto, CA, USA

**Keywords:** Cognitive Load, Emergency Medicine, Electronic Health Records, Wearable Sensors, Biomedical Informatics, Pilot Study

## Abstract

**Background:** Emergency Department (ED) physicians experience high cognitive workload that can impair patient safety and physician well-being. Despite this critical challenge, robust methods for real-time cognitive load measurement remain elusive. Cognitive Load Theory (CLT) provides a framework distinguishing intrinsic load (IL: inherent task complexity) from extraneous load (EL: factors that interfere with task completion). Building on this framework, the ALERT (Adaptive Load Estimation in Real-Time) project aims to develop dynamic cognitive load models using passively collected Electronic Health Record (EHR) data, validated through biometric and subjective measures.

**Objective:** This pilot study aimed to (1) demonstrate the feasibility of collecting, synchronizing, and processing multimodal data (EHR interactions, patient encounter data, wearable biometrics, subjective cognitive load reports) from ED physicians during clinical shifts, and (2) explore preliminary relationships between these data modalities to identify promising signals of cognitive load, including the utility of biometrics in predicting high subjective load.

**Methods:** Two ED physicians (Physician A: 11 shifts; Physician B: 7 shifts) from an academic medical center were monitored. Data included: (a) EHR interaction logs (15-minute aggregations) linked with (b) patient encounter data (ESI scores mapped to an inverted numeric acuity scale) to derive IL and EL proxies; (c) continuous wearable biometric data (EDA, HRV features processed in 5-minute windows, aggregated to 15 minutes); and (d) subjective cognitive load ratings (Paas Scale), entered when physicians experienced notable high or low load. Data were merged onto a synchronized 15-minute timeline. Pearson correlations explored relationships. Logistic regression using selected biometric features (LF/HF, heart rate, RMSSD, skin conductance level) plus a physician dummy variable was used to predict high subjective load (Paas *≥* 7).

**Results:** Multimodal data collection and synchronization were successfully achieved across 864 15-minute intervals. For Physician A, EHR activity diversity (EL proxy) showed a moderate positive correlation with Paas scores (r = 0.38), while maximum patient acuity (IL proxy) showed a smaller positive association (r = 0.10). For Physician B, several EL proxies demonstrated moderate positive correlations with Paas scores (r = 0.30–0.32), while the number of critical patients (IL proxy) showed a small positive association (r = 0.05). Physician A exhibited a moderate positive correlation between LF/HF ratio and Paas (r = 0.50). A logistic regression model using four selected biometric features and a physician identifier achieved an Area Under the ROC Curve (AUC) of 0.781 in discriminating high vs. low Paas scores (N = 60 observations with Paas data).

**Conclusion:** This pilot study demonstrates the feasibility of a multimodal approach for cognitive load assessment in the ED. Preliminary findings suggest EHR-derived load proxies correlate with subjective effort, and importantly, a small set of objective biometric features can discriminate high subjective cognitive load with good accuracy. These results strongly support the potential for the larger ALERT project.

## 1 Introduction

Emergency Department (ED) physicians work in one of the most cognitively demanding environments in healthcare [1–3]. They manage multiple, often high-acuity patients simultaneously [4], face frequent interruptions (exceeding 10 per hour [1, 5, 6]), and must make critical decisions under time pressure with incomplete information. This substantial cognitive workload can strain mental resources, leading to treatment delays, medical errors [7], and impaired diagnostic reasoning, ultimately affecting patient safety and contributing to fatigue [8–11]. Despite this critical link, robust, scalable and objective methods for measuring the dynamic cognitive burden on ED physicians in real time are largely absent [12].

Cognitive Load Theory (CLT) [13–15] provides a validated framework for characterizing the nature of cognitive challenges in such demanding settings. CLT distinguishes between: (1) *Intrinsic Load (IL)*, the inherent complexity of the clinical task itself (e.g., patient acuity [16]); (2) *Extraneous Load (EL)*, factors in the environment or interface that impede performance; and (3) *Germane Load (GL)*, the cognitive resources effectively allocated to learning and deep understanding. When the sum of IL and EL exceeds an individual’s finite cognitive capacity, cognitive overload occurs, diminishing resources available for germane processing, and impairing performance. EHR-based activity logs have emerged as a potential new source to measure aspects of this workload and its link to burnout [17–21]. Previous work has quantified time allocation in clinical settings [22], but passive EHR logs offer new scalability.

Current methods for assessing cognitive load, such as administrative metrics, direct observation, or subjective self-reports, have significant limitations [12]. Physiologic measurements, particularly heart rate variability (HRV), show promise as objective indicators [23–29]. The ALERT (Adaptive Load Estimation in Real-Time) project, for which this pilot study provides foundational work, proposes to address this gap by developing dynamic, CLT-based models of real-time cognitive load using passively collected EHR data [30], rigorously validated with physician biometrics and subjective effort assessments, building on concepts of multimodal monitoring [31].

This pilot study specifically aimed to:

1. Establish the technical feasibility of acquiring, synchronizing, and processing multimodal data streams [31]—including EHR interaction logs [17], patient encounter details, continuous physiological data from wearable sensors [32], and subjective cognitive load ratings—from ED physicians during their clinical shifts.

2. Conduct an exploratory analysis of these synchronized data to identify preliminary relationships between EHR-derived proxies of intrinsic and extraneous load, biometric stress indicators, and subjective reports of cognitive effort by physicians (Paas Scale [14]).

The findings of this pilot are intended to provide a proof of concept and inform the larger-scale development and validation efforts of the ALERT project. We hypothesized that even with limitations inherent in the pilot data, we would observe plausible associations consistent with CLT, thus supporting the utility of our proposed multimodal approach.

## 2 Methods

### 2.1 Study Design and Participants

This pilot study used a prospective observational design involving two board-certified Emergency Department (ED) physicians working at Stanford University Hospital, a suburban, academic, Level I trauma center with over 100,000 annual patient visits. The study was conducted over a 4-week period in May 2025. Physician A participated for 11 clinical shifts and physician B for 7 clinical shifts; Both physicians worked a variety of shifts that differed in patient acuity, trainee supervision responsibilities, and time of day.

### 2.2 Data Acquisition

Multimodal data streams were collected and synchronized as follows:

#### 2.2.1 EHR Interaction and Patient Encounter Data

Patient information and physician’s EHR actions measured by the EHR audit log were extracted from the institution’s Epic EHR system (Epic Systems Corporation, Verona, WI). The raw data provided hourly summaries of physician activity, including patient encounter identifiers for patient-specific interactions, type of EHR task, and seconds of EHR activity in each 15-minute segment..

Patient encounter details were also extracted for all cases where participating physicians documented interactions in the audit log. These details included patient age, arrival information, Emergency Severity Index (ESI) triage acuity scores, and the number of documented chief complaints and existing diagnoses.

#### 2.2.2 Wearable Biometric Data

Continuous physiological data were collected using an Empatica EmbracePlus (Empatica Inc., Milan, Italy) research-grade wearable device worn by the physicians throughout their monitored shifts. The device captured:

- Photoplethysmography (PPG) at 64 Hz, used for deriving heart rate (HR) and heart rate variability (HRV) metrics
- Electrodermal activity (EDA) at 4 Hz, used for skin conductance level (SCL) and skin conductance responses (SCRs)
- Skin temperature at 1 Hz
- 3-axis accelerometry (ACC) at 31.25 Hz

Raw data were processed using a custom Python-based pipeline (including modules for AVRO parsing, data cleaning, and feature extraction, referencing tools such as *NeuroKit2* and *tsfresh* as appropriate). Key biometric features were extracted over 5-minute rolling windows, including time-domain HRV (RMSSD, MeanNN, SDNN), frequency-domain HRV (LF, HF, LF/HF ratio), EDA features (SCL mean, SCR count), and mean heart rate.

#### 2.2.3 Subjective Cognitive Load Data

Subjective cognitive load was assessed using the Paas Scale [14], a single-item 9-point Likert scale rating perceived mental effort for the immediately preceding cognitive task or period. Physicians were instructed to enter Paas scores via a smartphone application whenever they experienced particularly high or low cognitive load periods. Free-text comments associated with Paas ratings were also collected.

#### 2.2.4 Shift Schedule Data

Physician shift schedules (start and end times) were obtained to define on-shift periods for analysis.

### 2.3 Data Processing and Synchronization

1. **EHR Data Processing (15-minute resolution):** The quarter-hourly EHR interaction data were unpivoted to create a row for each 15-minute interval containing EHR activity. The *activity hour utc dttm* was used to define the start of each hour, and 15-minute offsets were applied to establish *quarter start time* for each interval.
  - **Extraneous Load (EL) Proxies:** For each physician and 15-minute quarter, the following EL proxies were calculated: *patient count* (the number of unique patient charts accessed), *task count* (the number of distinct EHR tasks performed), *total time* (the number of seconds active in the quarter), *task switches* (the number of switches in EHR task type), and *patient switches* (the number of switches between different patient charts).
  - **Intrinsic Load (IL) Proxies:** Patient encounter data were merged with the 15-minute EHR interaction data using unique patient identifiers. The ESI score was converted to a numeric scale where ESI 1 (highest acuity) was mapped to 5, ESI 2 to 4, and so forth, with ESI 5 (lowest acuity) mapped to 1. IL proxies such as the maximum patient acuity were then derived using this inverted scale, so higher IL proxy values reflect higher actual patient acuity. For each physician and 15-minute quarter containing patient interactions, the following IL proxies were calculated based on the patients accessed in that quarter: *average acuity* (the average acuity score of patients the physician interacted with during the quarter), *maximum acuity* (the highest single acuity score of the patients that the physician interacted with during the quarter), *total acuity* (the sum of all patient acuities the physician interacted with during the quarter), *chief complaint count* (the average number of chief complaints for patients the physician interacted with during the quarter), *diagnosis count* (the average number of existing diagnoses patients had that the physician interacted with during the quarter), and *critical patients* (the count of patients with ESI *≤* 2 that the physician interacted with during the quarter).
2. **Biometric Data Aggregation:** The 5-minute windowed biometric features were aggregated to 15-minute means, aligned with the quarter start times of the EHR data.
3. **Subjective Load Data Alignment:** Paas scores were aligned to the quarter time interval they most likely corresponded to (typically the interval immediately preceding the Paas entry timestamp). If multiple Paas scores fell into a single 15-minute window, their mean was taken.
4. **Master Timeline Creation:** A master timeline was constructed for each physician, covering all their recorded shifts with 15-minute intervals. The processed EL proxies, IL proxies, aggregated biometric features, and aligned Paas scores were merged onto this timeline. Intervals with no EHR activity had EL/IL counts set to zero; average-based IL metrics were kept as NaN if no patient interactions occurred in a window. Biometric and Paas data remained NaN if no data were available for a given 15-minute window.

### 2.4 Statistical Analysis

All analyses were conducted using Python (version 3.13) with pandas, *NumPy, SciPy*, and *statsmodels/scikit-learn* libraries. Exploratory analysis focused on Pearson correlation coefficients (r) to assess linear relationships between:

- EHR-derived IL proxies and individual biometric features (RMSSD, MeanNN, SDNN, LF, HF, LF/HF, SCL mean, SCR count, hr mean)
- EHR-derived EL proxies and individual biometric features
- EHR-derived IL/EL proxies and Paas scores
- Individual biometric features and Paas scores

Correlations were examined separately for each physician due to the small sample size and potential for individual differences.

To explore the combined utility of features in predicting subjective load, Paas scores were dichotomized (Paas *≥* 7 indicating high load, Paas *<* 7 indicating low load). Logistic regression models were then trained using a selected set of four biometric features (LF/HF, heart rate, RMSSD, and mean skin conductance level) plus a physician dummy variable (Physician B vs. Physician A as reference) to predict high vs. low Paas. Model performance was assessed using the Area Under the Receiver Operating Characteristic curve (AUC). An attempt to fit a more comprehensive model using all EHR and biometric features was also made. Given the exploratory nature of this pilot study with N = 2, p-values for correlations were not the primary focus; rather, the magnitude and direction of correlations were examined for consistency with CLT and to identify promising signals for future investigation.

## 3 Results

### 3.1 Data Overview

Data were collected from Physician A over 11 shifts and Physician B over 7 shifts, resulting in a total of 864 analyzable 15-minute on-shift intervals after processing and synchronization. Subjective Paas scores were available for 60 of these 15-minute intervals, which formed the basis for predictive modeling of high cognitive load. An illustrative example of the synchronized multimodal data for Physician B over an 8-hour shift segment is shown in Figure 1.

**Figure 1.**
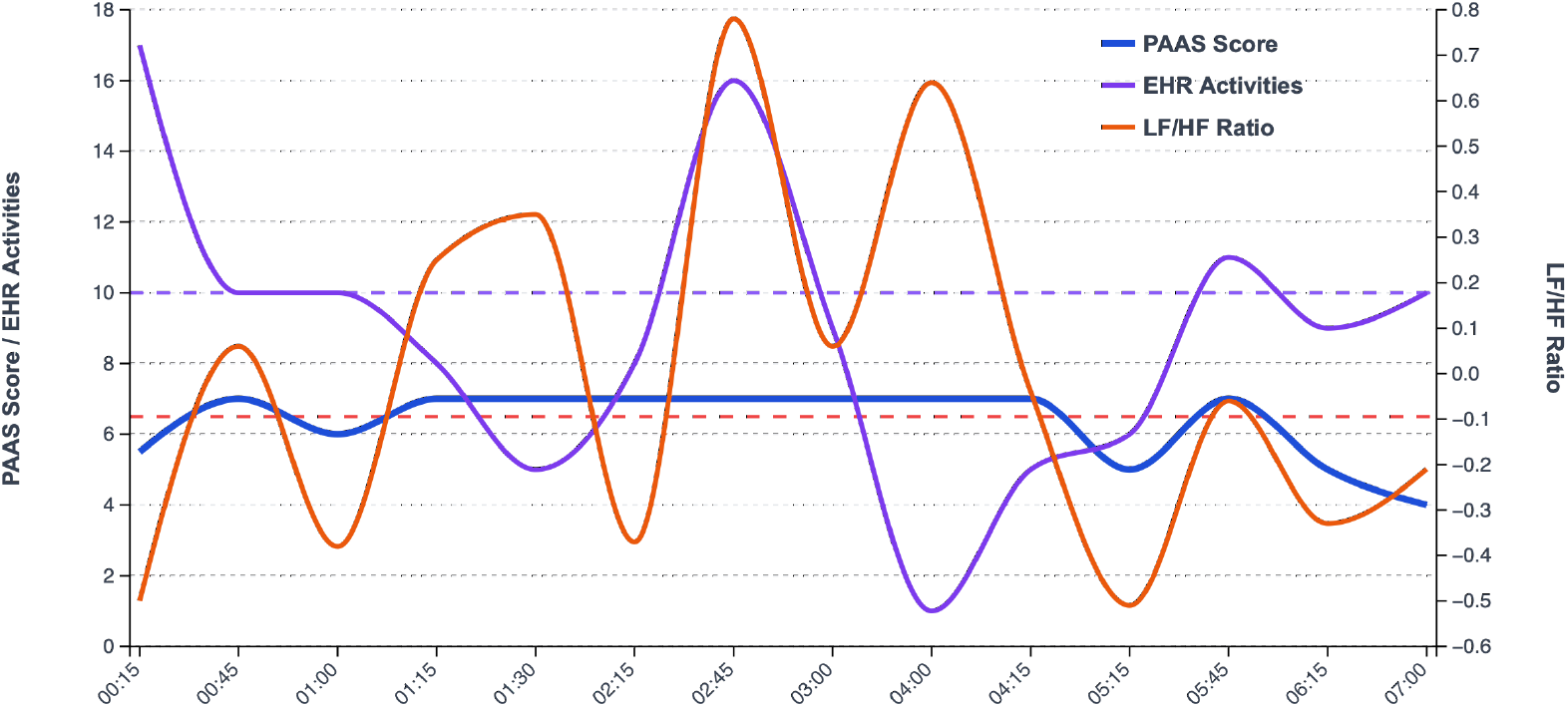
Illustrative time series for Physician B during an 8-hour shift segment on May 10, 2025. Shown are subjective Paas Scores (blue), EHR unique activities per 15-min interval (purple), and LF/HF ratio (orange). Dashed horizontal lines indicate thresholds for high stress (Paas *≥* 6.5, red) and high EHR workload (*≥* 10 activities, purple). Correlation for this segment: Paas vs. EHR Activities r = 0.321; Paas vs. LF/HF Ratio r = 0.220 (N = 14 intervals).

### 3.2 Correlations with Subjective Cognitive Load (Paas Scale)

Pearson correlations were computed separately for each physician to explore relationships between EHR-derived load proxies, biometric features, and Paas scores.

#### 3.2.1 EHR-Derived Load Proxies vs. Paas Scores

For Physician A, *task count* (an EL proxy) demonstrated a moderate positive correlation with Paas scores (r = 0.38, N = 22 data points with Paas). The IL proxy *chief complaint count* also showed a positive correlation (r = 0.26), while *maximum acuity* (inverted ESI, higher value = higher acuity) had a smaller positive correlation (r = 0.10).

For Physician B, several EL proxies showed moderate positive correlations with Paas scores: *activity count* (r = 0.32), *total time* (r = 0.30), and *task switches* (r = 0.31). The IL proxy *total acuity* (r = 0.26) showed a notable positive association. Detailed correlations are presented in Tables 2 and 3.

**Table 1:** Empatica EmbracePlus Data Streams, Derived Features, and Clinical Relevance

| Data Stream | Sampling Rate | Raw Elements | Derived Feature | Units | Definition & Clinical Relevance |
| --- | --- | --- | --- | --- | --- |
| **Photoplethysmography (PPG)** | 64 Hz | Inter-beat intervals, Peak detection timestamps | RMSSD | ms | Root mean square of successive RR interval differences. Reflects parasympathetic activity; decreases with stress. |
|  |  |  | MeanNN | ms | Mean of normal RR intervals. Average time between heartbeats. |
|  |  |  | SDNN | ms | Standard deviation of RR intervals. Overall autonomic activity measure. |
|  |  |  | LF Power | ms<sup>2</sup> | Low-frequency power (0.04–0.15 Hz). Mixed sympathetic and parasympathetic influences. |
|  |  |  | HF Power | ms<sup>2</sup> | High-frequency power (0.15–0.4 Hz). Primarily reflects parasympathetic activity. |
|  |  |  | LF/HF Ratio | – | Sympathovagal balance ratio. Higher values suggest stress or sympathetic dominance. |
|  |  |  | Heart Rate Mean | bpm | Average heart rate over window. Complex relationship with cognitive load in clinical settings. |
| **Electrodermal Activity (EDA)** | 4 Hz | Skin conductance amplitude (μS) | SCL mean | μS | Mean skin conductance level. Reflects tonic arousal; increases with cognitive load and stress. |
|  |  |  | SCR count | peaks/5 min | Number of skin conductance responses. Reflects phasic arousal events; acute stress indicator. |
| **Skin Temperature** | 1 Hz | Temperature readings | Temp mean | °C | Mean skin temperature over window. Can reflect autonomic changes and stress response patterns. |
| **3-Axis Accelerometry** | 31.25 Hz | X, Y, Z acceleration vectors | Activity mean | g | Mean activity level over window. Used to control for movement artifacts and physical activity confounds. |

**Table 2:** Correlations (Pearson r) between EHR-Derived Load Proxies and Paas Scores for Physician A (N = 11 shifts)

| **EHR-Derived Load Proxy** | **Correlation with Paas Score ($r$)** |
| --- | --- |
| *Intrinsic Load (IL) Proxies* |  |
| Average Acuity (Inverted ESI) | 0.17 |
| Maximum Acuity (Inverted ESI) | 0.10 |
| Total Acuity (Inverted ESI) | 0.09 |
| Chief Complaint Count | 0.26 |
| Diagnoses Count | -0.24 |
| Critical Patients (ESI 1–2) | -0.03 |
| *Extraneous Load (EL) Proxies* |  |
| Patient Count | -0.02 |
| Activity Count | 0.38 |
| Total Time | -0.04 |
| Task Switches | 0.24 |
| Patient Switches | 0.02 |

**Table 3:** Correlations (Pearson r) between EHR-Derived Load Proxies and Paas Scores for Physician B (N = 7 shifts)

| **EHR-Derived Load Proxy** | **Correlation with Paas Score ($r$)** |
| --- | --- |
| *Intrinsic Load (IL) Proxies* |  |
| Average Acuity (Inverted ESI) | 0.03 |
| Maximum Acuity (Inverted ESI) | 0.09 |
| Total Acuity (Inverted ESI) | 0.26 |
| Chief Complaint Count | 0.15 |
| Diagnoses Count | -0.12 |
| Critical Patients (ESI 1–2) | 0.05 |
| *Extraneous Load (EL) Proxies* |  |
| Patient Count | 0.17 |
| Activity Count | 0.32 |
| Total Time | 0.30 |
| Task Switches | 0.31 |
| Patient Switches | 0.24 |

#### 3.2.2 Biometric Features vs. Paas Scores

Physician A exhibited a strong positive correlation between the LF/HF ratio and Paas scores (r = 0.50, N = 9 data points with Paas), and a moderate negative correlation between mean heart rate and Paas (r = *−*0.48).

For Physician B, correlations between biometric features and Paas scores were generally weaker. MeanNN interval showed a small negative correlation (r = *−*0.21), as did RMSSD (r = *−*0.18). The LF/HF ratio showed a small positive correlation with Paas (r = 0.22). Selected biometric-Paas correlations are presented in Table 4.

**Table 4:**
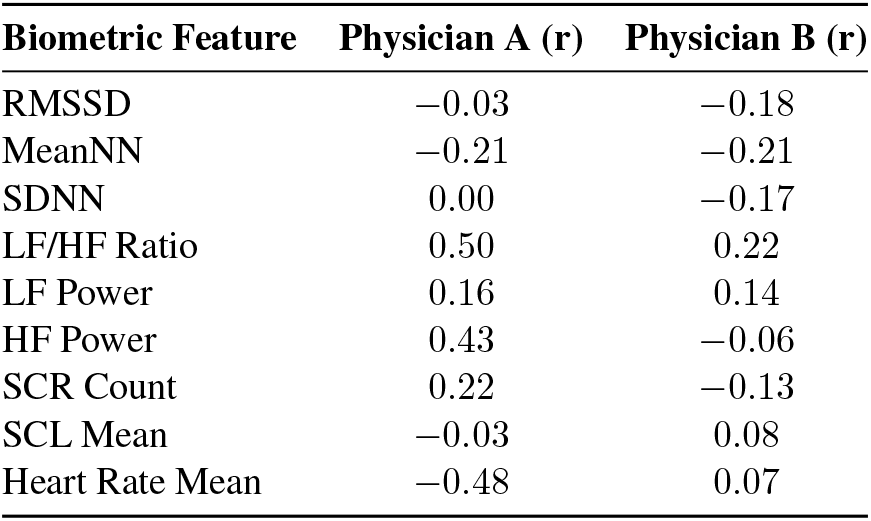
Correlations (Pearson r) between Selected Biometric Features and Paas Scores

### 3.3 Correlations between EHR-Derived Load Proxies and Biometrics

Associations between EHR-derived load and biometric features were explored. For instance, for Physician B, average patient acuity (IL proxy) showed a moderate positive correlation with normalized mean heart rate (r = 0.30, N = 98 data points). Other correlations were generally weaker and varied between physicians.

### 3.4 Predicting High Subjective Cognitive Load

A logistic regression model was developed to predict high subjective cognitive load (Paas score *≥* 7) using a selected set of four biometric features (LF/HF ratio, Heart Rate Mean, RMSSD, SCL Mean) and a physician identifier (Physician B vs. Physician A as reference), based on N = 60 intervals with Paas data. The model achieved an Area Under the ROC Curve (AUC) of 0.781 (Figure 2).

**Figure 2.**
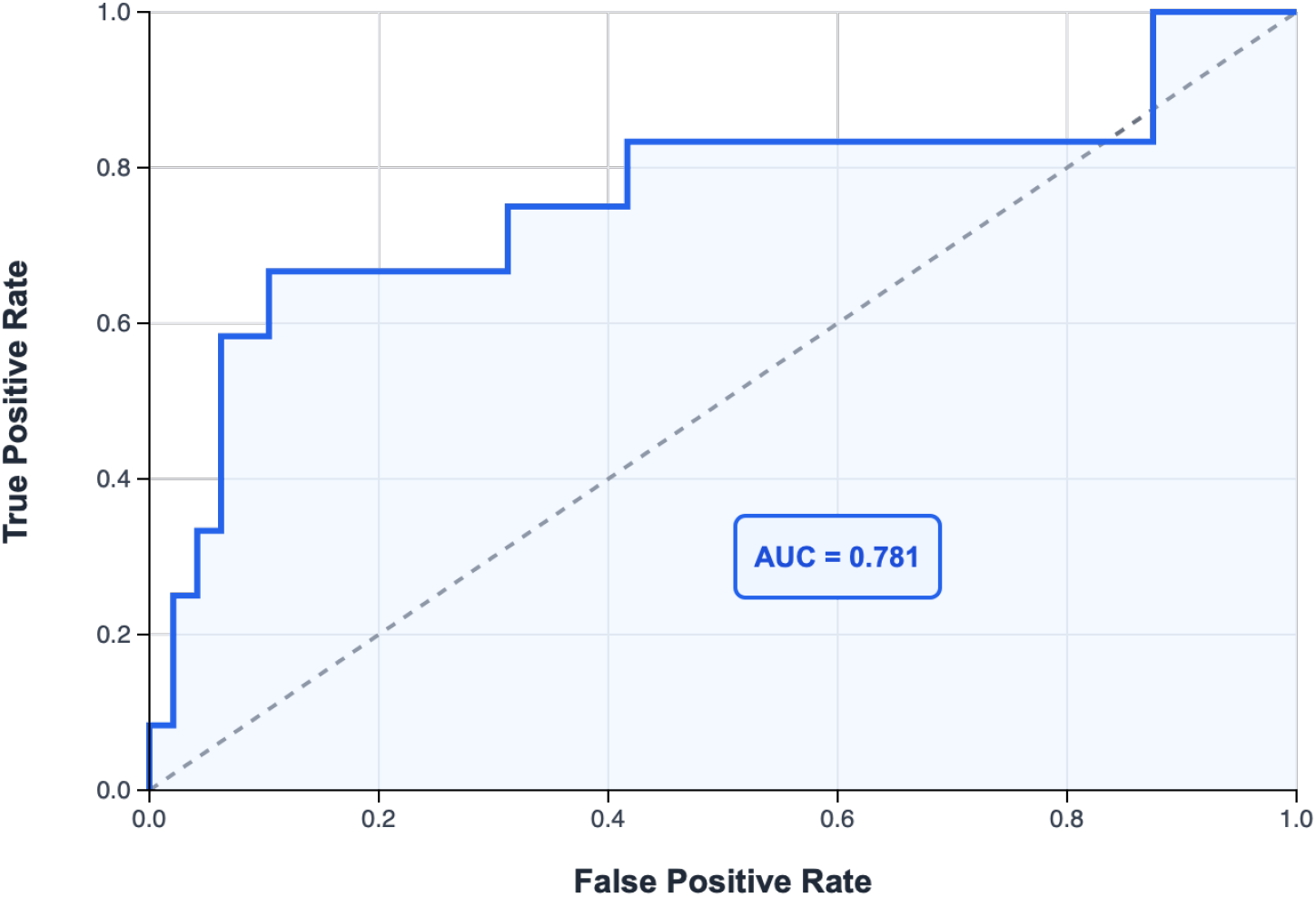
ROC Curve for Logistic Regression Model Predicting High Subjective Cognitive Load (Paas Score *≥* 7) using Selected Biometric Features (LF/HF Ratio, Heart Rate Mean, RMSSD, SCL Mean) and a Physician Indicator. Model performance yielded an AUC of 0.781 (N = 60 observations with Paas data).

**Figure 3.**
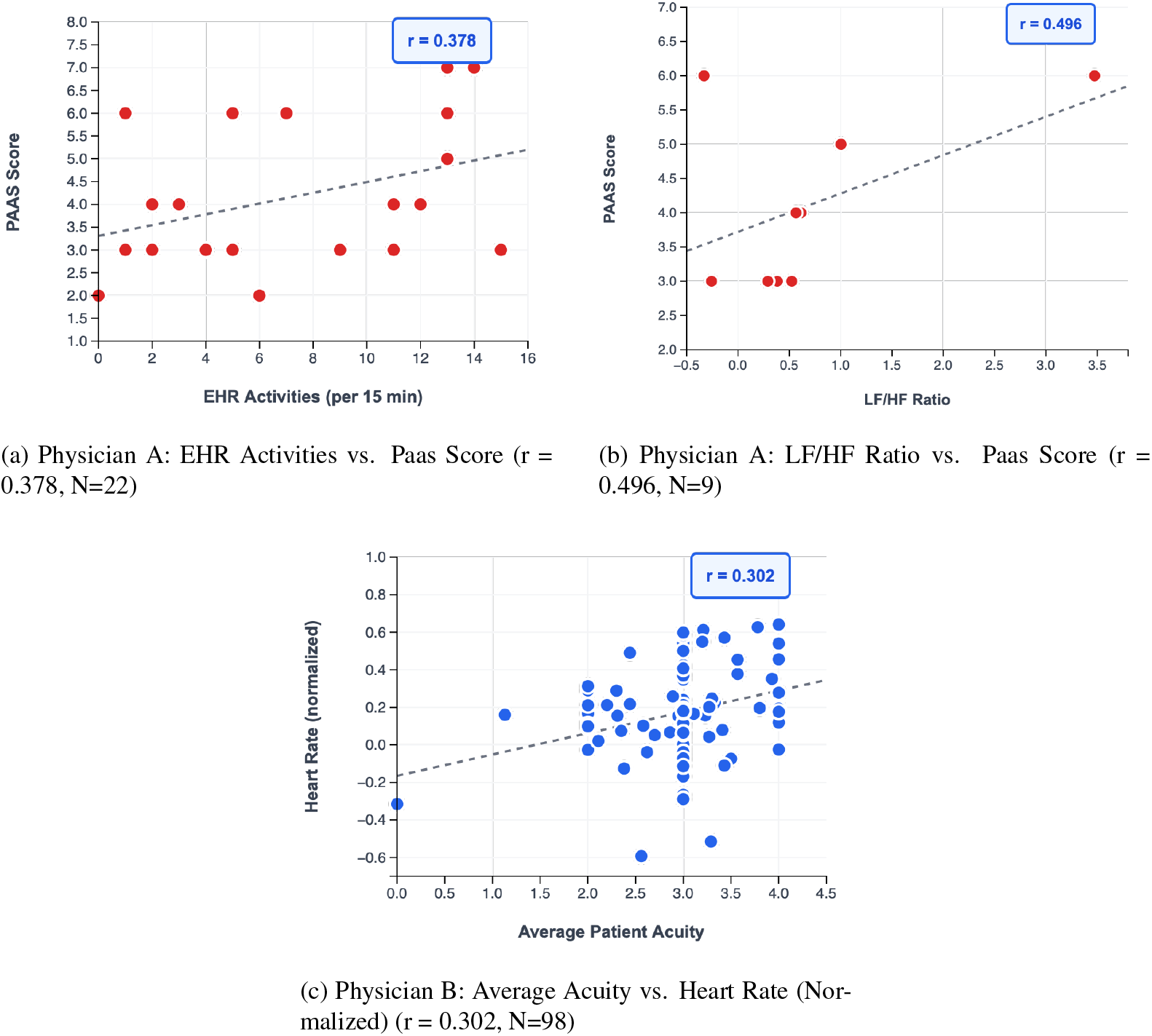
Scatter plots illustrating key correlations. (A) Relationship between activity count (EL proxy) and subjective Paas scores for Physician A. (B) Relationship between LF/HF ratio and subjective Paas scores for Physician A. (C) Relationship between average acuity (IL proxy; higher value = higher acuity) and normalized mean heart rate for Physician B. Each point represents a 15-minute interval; dashed lines indicate linear regression fit. N values represent the number of data points available for each specific correlation.

The model coefficients are presented in Table 5; RMSSD was a statistically significant predictor (p = 0.032). An attempt to fit a more comprehensive logistic regression model using all available EHR-derived IL/EL proxies and all biometric features was unsuccessful due to matrix singularity, likely due to the limited sample size relative to the number of predictors.

**Table 5:**
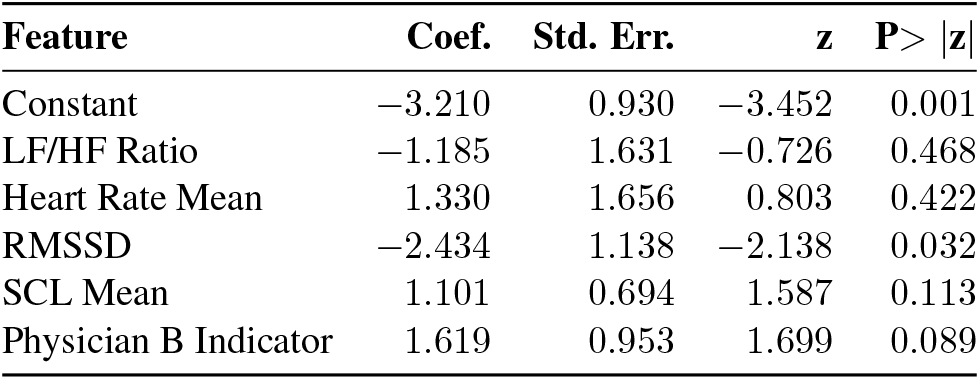
Logistic Regression Coefficients for Predicting High Paas Score (*≥* 7) using Selected Biometric Features (N = 60)

## 4 Discussion

This pilot study established the feasibility of a multimodal approach to assess cognitive load in Emergency Department (ED) physicians. We successfully collected, synchronized, and analyzed EHR interactions, patient encounter data, wearable biometrics, and subjective cognitive load reports during clinical shifts. The generation of a synchronized 15-minute interval dataset from 864 observations provided a platform to explore relationships between EHR-derived proxies for intrinsic load (IL) and extraneous load (EL), physicians’ subjective effort (Paas Scale), and physiological responses, supporting the ALERT project’s objectives.

Consistent with Cognitive Load Theory [13, 14], EHR-derived load proxies showed positive associations with physicians’ subjective Paas scores. Specifically, for Physician A, increased EHR activity diversity (an EL proxy reflecting more varied tasks) correlated notably with higher Paas scores (r = 0.38). For Physician B, multiple EL proxies, including EHR activity diversity (r = 0.32), total EHR active time (r = 0.30), and task switches (r = 0.31), demonstrated moderate correlations with increased subjective effort. These observations align with existing literature indicating that increased task switching and system interaction complexity contribute to cognitive burden [8, 19, 33, 34].

Furthermore, IL proxies, such as patient acuity (for Physician A, r = 0.10 for maximum acuity; for Physician B, r = 0.26 for total acuity) and average number of chief complaints (Physician A, r = 0.26), also showed positive correlations with Paas scores. This supports the premise that greater inherent task complexity increases perceived effort.

Analysis of biometric features against subjective Paas scores revealed individual differences and underscored the utility of physiological monitoring [23, 26]. Physician A exhibited a strong positive correlation between LF/HF ratio and Paas scores (r = 0.50), indicative of increased sympathetic nervous system activity (or reduced parasympathetic modulation) during periods of higher perceived effort; this is consistent with established physiological stress responses. Conversely, Physician A also showed a negative correlation between mean heart rate and Paas scores (r = *−*0.48). This latter finding may reflect periods of focused attention or specific coping mechanisms, rather than a simple linear stress response.

For Physician B, biometric correlations with Paas scores were generally weaker. However, trends such as negative associations with RMSSD (r = *−*0.18) and MeanNN (r = *−*0.21) suggested reduced vagal tone accompanying higher effort. The feasibility of using smartwatch physiological signals for stress prediction, as demonstrated by studies like Dai et al. (2021) [24], lends support to the validity of our biometric approach.

A key outcome of this pilot study was the successful discrimination between high (Paas *≥* 7) and low subjective cognitive load. A logistic regression model, utilizing four biometric features (LF/HF ratio, mean heart rate, RMSSD, SCL mean) and a physician identifier (Physician B vs. Physician A as reference), achieved an Area Under the ROC Curve (AUC) of 0.781. This performance is particularly encouraging given the small sample size (N=2 physicians) and demonstrates the potential for objective, biometric-based assessment of cognitive load. Notably, RMSSD emerged as a statistically significant predictor in this model (p = 0.032), reinforcing the value of HRV metrics in objectively assessing cognitive states [23]. This provides strong support for the predictive modeling aims of the ALERT project.

Correlations between EHR-derived load proxies and biometric features were more varied and generally weaker than those observed with Paas scores. For instance, Physician B demonstrated a moderate positive correlation between average patient acuity (an IL proxy) and mean heart rate (r = 0.30). These mixed results likely reflect the complexity of physiological responses to workload, the potential influence of the 15-minute data aggregation window, and the importance of accounting for individual differences in stress response patterns.

### 4.1 Limitations

This pilot study has several important limitations that must be acknowledged. First, the sample size (N = 2 physicians) is very small, precluding generalization and limiting statistical power. The findings should be interpreted as exploratory and hypothesis-generating rather than definitive. Second, EHR interaction data were aggregated at 15-minute intervals, which may mask finer-grained temporal dynamics of cognitive load fluctuations [17, 18].

Third, Paas scores were collected on an ad libitum basis when physicians felt they were experiencing particularly high or low cognitive load, which might introduce selection bias and limit the representativeness of the cognitive load spectrum [12, 14]. Physicians may not have had time to perform the Paas scale when truly overloaded, limiting more granular understanding of this relationship. A more systematic sampling approach would strengthen future studies. Fourth, the comprehensive logistic regression model failed due to matrix singularity, highlighting the challenge of working with limited sample sizes relative to the number of potential predictors.

Finally, this pilot did not include detailed contextual observations of interruptions [1, 33], patient flow patterns, or other environmental factors that are known to influence cognitive load in the ED [2]. The individual differences observed between the two physicians also suggest that personalized models may be necessary for optimal performance.

### 4.2 Future Directions

The pilot study’s AUC of 0.781 for predicting high cognitive load validates the multimodal approach and supports the progression of the ALERT project. Immediate future work will focus on expanding the participant cohort to include a more diverse sample of ED physicians (differing experience levels, shift types, and clinical environments) to enhance generalizability. The methodological framework established herein provides a robust foundation for this larger-scale data collection.

Subsequent research will address technical enhancements. These include increasing the temporal resolution of data acquisition, implementing more advanced feature engineering techniques, and integrating supplementary contextual data sources, such as patient flow metrics and departmental census information. To improve predictive performance beyond the baseline logistic regression model, advanced machine learning approaches, including ensemble methods and deep learning architectures, will be systematically evaluated.

The development of real-time cognitive load monitoring systems presents distinct implementation challenges that will be addressed. These include ensuring data privacy, facilitating seamless integration into physician workflow, and designing effective, actionable feedback mechanisms. The primary objective remains the creation of systems capable of providing early warnings for cognitive overload and suggesting targeted interventions to optimize physician performance and well-being.

## 5 Conclusion

This pilot study successfully demonstrates the feasibility of a multimodal approach to capture dynamic data streams relevant to understanding ED physician cognitive load. Despite the inherent limitations of a small-scale pilot study, the preliminary findings are encouraging and provide strong evidence for the viability of the ALERT project approach.

Key achievements include: (1) successful multimodal data collection and synchronization across 864 15-minute intervals; (2) identification of plausible correlations between EHR-derived proxies for intrinsic and extraneous load and physicians’ subjective reports of cognitive effort; and (3) demonstration that a focused model utilizing objective wearable biometric features can discriminate periods of high subjective cognitive load with good accuracy (AUC = 0.781).

These results provide a solid foundation for the larger, more comprehensive ALERT project and support the potential for developing real-time, objective cognitive load assessment tools that could ultimately improve both physician well-being and patient safety in emergency medicine. The multimodal approach validated here represents a significant step toward more sophisticated understanding and management of cognitive burden in high-stakes clinical environments.

## Data Availability

All data produced in the present study are available upon reasonable request to the authors.

## References

[1] Chisholm CD, Collison EK, Nelson DR, Cordell WH. Emergency Department Workplace Interruptions: Are Emergency Physicians “Interrupt-driven” and “Multitasking”? Acad Emerg Med. 2000;7(11):1239–1243.

[2] Patel VL, Kannampallil TG, Shortliffe EH. Role of cognition in generating and mitigating clinical errors. BMJ Qual Saf. 2015;24(7):468–474.

[3] Horsky J, Zhang J, Patel VL. To err is not entirely human: complex technology and user cognition. J Biomed Inform. 2005;38(4):264–266.

[4] Laxmisan A, Hakimzada F, Sayan OR, Green RA, Zhang J, Patel VL. The multitasking clinician: decision-making and cognitive demand during and after team handoffs in emergency care. Int J Med Inform. 2007;76(11–12):801–811.

[5] El-Sherif N, Hawthorne HJ, Forsyth KL, Abdelrahman A, Hallbeck SM, Blocker RC. Physician interruptions and workload during Emergency Department shifts. Proc Hum Factors Ergon Soc Annu Meet. 2017;61(1):649–652.

[6] Kissler MJ, Kissler K, Burden M. Toward a medical “ecology of attention.” N Engl J Med. 2021;384(4):299–301.

[7] Croskerry P, Sinclair D. Emergency medicine: A practice prone to error? CJEM. 2001;3(4):271– 276.

[8] Melnick ER, Harry E, Sinsky CA, Dyrbye LN, Wang H, Trockel MT, West CP, Shanafelt T. Perceived Electronic Health Record Usability as a Predictor of Task Load and Burnout Among US Physicians: Mediation Analysis. J Med Internet Res. 2020;22(12):e23382.

[9] Franklin BD, Birch S, Savage I, Wong M, Woloshynowych M, Jacklin A, Barber N. Methodological variability in detecting prescribing errors and consequences for the evaluation of interventions. Pharmacoepidemiol Drug Saf. 2009;18(11):992–999.

[10] Khairat S, Coleman C, Ottmar P, Jayachander DI, Bice T, Carson SS. Association of electronic health record use with physician fatigue and efficiency. JAMA Netw Open. 2020;3(6):e207385.

[11] Gardner RL, Cooper E, Haskell J, et al. Physician stress and burnout: the impact of health information technology. J Am Med Inform Assoc. 2019;26(2):106–114.

[12] Vella KM, Hall AK, van Merrienboer JJG, Hopman WM, Szulewski A. An exploratory investigation of the measurement of cognitive load on shift: Application of cognitive load theory in emergency medicine. AEM Educ Train. 2021;5(4):e10634.

[13] Sweller J. Cognitive Load Theory. In: Ross BH, editor. Psychology of Learning and Motivation. Vol. 55. Elsevier Academic Press; 2011. p. 37–76.

[14] Paas Fgwc. Training strategies for attaining transfer of problem-solving skill in statistics: A cognitive-load approach. J Educ Psychol. 1992;84(4):429–434.

[15] Paas F, Renkl A, Sweller J. Cognitive load theory and instructional design: Recent developments. Educational Psychologist. 2003;38(1):1–4.

[16] Held N, Neumeier A, Amass T, et al. Extraneous load, patient census, and patient acuity correlate with cognitive load during ICU rounds. Chest. 2024;165(6):1448–1457.

[17] Adler-Milstein J, Adelman JS, Tai-Seale M, Patel VL, Dymek C. EHR audit logs: A new goldmine for health services research? J Biomed Inform. 2020;101:103343.

[18] Sinsky CA, Rule A, Cohen G, Arndt BG, Shanafelt TD, Sharp CD, Baxter SL, Tai-Seale M, Yan S, Chen Y, Adler-Milstein J, Hribar M. Metrics for assessing physician activity using electronic health record log data. J Am Med Inform Assoc. 2020;27(4):639–643.

[19] Moy AJ, Cato KD, Kim EY, Withall J, Rossetti SC. A computational framework to evaluate emergency department clinician task switching in the electronic health record using event logs. AMIA Annu Symp Proc. 2023;2023:1183–1192.

[20] Rose C, Thombley R, Noshad M, et al. Team is brain: leveraging EHR audit log data for new insights into acute care processes. J Am Med Inform Assoc. 2022;30(1):8–15.

[21] Kannampallil T, Abraham J, Lou SS, Payne PRO. Conceptual considerations for using EHR-based activity logs to measure clinician burnout and its effects. J Am Med Inform Assoc. 2021;28(5):1032–1037.

[22] Sinsky C, Colligan L, Li L, et al. Allocation of physician time in ambulatory practice: A time and motion study in 4 specialties. Ann Intern Med. 2016;165(11):753–760.

[23] Mullikin DR, Flanagan RP, Merkebu J, Durning SJ, Soh M. Physiologic measurements of cognitive load in clinical reasoning. Diagnosis. 2024;11(2):125–131.

[24] Dai R, Lu C, Yun L, Lenze E, Avidan M, Kannampallil T. Comparing stress prediction models using smartwatch physiological signals and participant self-reports. Comput Methods Programs Biomed. 2021;208:106207.

[25] Sundrani S, Chen J, Jin BT, Shakeri Hossein Abad Z, Rajpurkar P, Kim D. Predicting patient decompensation from continuous physiologic monitoring in the emergency department. npj Digit Med. 2023;6(1):60.

[26] Johannessen E, Szulewski A, Radulovic N, White M, Braund H, Howes D, Rodenburg D, Davies C. Psychophysiologic measures of cognitive load in physician team leaders during trauma resuscitation. Comput Human Behav. 2020;111:106393.

[27] Kim D, Jin BT. Development and comparative performance of physiologic monitoring strategies in the emergency department. JAMA Netw Open. 2022;5(9):e2233712.

[28] Szulewski A, Gegenfurtner A, Howes DW, Sivilotti MLA, van Merriënboer JJG. Measuring physician cognitive load: validity evidence for a physiologic and a psychometric tool. Adv Health Sci Educ Theory Pract. 2017;22(4):951–968.

[29] Park SH, Goldberg SA, Al-Ballaa A, et al. Objective measurement of learners’ cognitive load during simulation-based trauma team training: A pilot study. J Surg Res. 2022;279:361–367.

[30] Kim S, Warner BC, Lew D, Lou SS, Kannampallil T. Measuring cognitive effort using tabular transformer-based language models of electronic health record-based audit log action sequences. J Am Med Inform Assoc. 2024;31(10):2228–2235. doi:10.1093/jamia/ocae171.

[31] Kansal A, Chen E, Jin T, Rajpurkar P, Kim D. Multimodal Clinical Monitoring in the Emergency Department (MC-MED). Published online March 3, 2025. doi:10.13026/JZ99-4J81.

[32] Agarwal AK, Gonzales R, Scott K, Merchant R. Investigating the Feasibility of Using a Wearable Device to Measure Physiologic Health Data in Emergency Nurses and Residents: Observational Cohort Study. JMIR Form Res. 2024;8:e51569.

[33] Westbrook JI, Raban MZ, Walter SR, Douglas H. Task errors by emergency physicians are associated with interruptions, multitasking, fatigue and working memory capacity: a prospective, direct observation study. BMJ Qual Saf. 2018;27(8):655–663.

[34] Morra M, Braund H, Hall AK, Szulewski A. Cognitive load and processes during chest radiograph interpretation in the emergency department across the spectrum of expertise. AEM Educ Train. 2021;5(4):e10693.

